# Causality between Diabetes and MN: Mendelian randomization and meta-analysis

**DOI:** 10.1101/2024.04.25.24306363

**Authors:** Zhihang Su, Ziqi Luo, Di Wu, Wen Liu, Wangyang Li, Zheng Yin, Rui Xue, Liling Wu, Yuan Cheng, Qijun Wan

**Author notes:** **Correspondence:** Corresponding Author: Qijun Wan.

## Abstract

**Background:** Membranous nephropathy (MN) has not yet been fully elucidated regarding its relationship with Type I and II Diabetes. This study aims to evaluate the causal effect of multiple types of diabetes and MN by summarizing the evidence from the Mendelian randomization (MR) study.

**Methods:** The statistical data for MN was obtained from a GWAS study encompassing 7,979 individuals. Regarding diabetes, fasting glucose, fasting insulin, and HbA1C data, we accessed the UK-Biobank, within family GWAS consortium, MAGIC, Finnish database, MRC-IEU, and Neale Lab, which provided sample sizes ranging from 17,724 to 298,957. As a primary method in this MR analysis, we employed the Inverse Variance Weighted (IVW), Weighted Median, Weighted mode, MR-Egger, Mendelian randomization pleiotropy residual sum, and outlier (MR-PRESSO) and Leave-one-out sensitivity test. Reverse MR analysis was utilized to investigate whether MN affects Diabetes. Meta-analysis was applied to combine study-specific estimates.

**Results:** It has been determined that type 2 diabetes, gestational diabetes, type 1 diabetes with or without complications, maternal diabetes, and insulin use pose a risk to MN. Based on the genetic prediction, fasting insulin, fasting blood glucose, and HbA1c levels were not associated with the risk of MN. No heterogeneity, horizontal pleiotropy, or reverse causal relationships were found. The meta-analysis results further validated the accuracy.

**Conclusions:** The MR analysis revealed the association between MN and various subtypes of diabetes. This study has provided a deeper understanding of the pathogenic mechanisms connecting MN and diabetes.

## 1 Introduction

Membranous nephropathy (MN), the most common etiology in adult nephrotic syndrome, is an immune-related disease(1). Approximately 80% of patients cannot identify a specific cause and are referred to as primary MN. The formation of immune complexes in the mesangial area, typically comprising immunoglobulin G (IgG), associated antigens, and complement components, including the membrane attack complex (MAC), results in substantial thickening of the glomerular capillary walls(2). Immune dysregulation resulting from this immune conflict disrupts the structural integrity of podocytes, leading to significant proteinuria. Spontaneous complete remission rates of untreated MN have been reported to range from 20% to 30%, and advancements have been achieved in utilising immunosuppressive drugs to manage MN. However, around 10% of patients with MN eventually develop end-stage renal disease (ESRD), and within a 10-year timeframe, 40-50% of individuals with nephrotic syndrome encounter kidney failure.

Diabetes is an increasingly severe global public health issue, with its prevalence increasing year by year. As of 2019, approximately 463 million adults (one in ten) worldwide have diabetes, of which half remain undiagnosed(3). In addition, diabetes in young people is also on the rise. Type 1 diabetes in childhood and type 2 diabetes in adolescence are both increasing. The medical costs associated with diabetes and its complications are incalculable(4). Diabetic nephropathy (DN) is a common complication of diabetes. It has become a significant cause of chronic kidney disease (CKD) and end-stage renal disease (ESRD) with the increasing prevalence of diabetes(5). In the past, the diagnosis of diabetic nephropathy mainly relied on clinical diagnosis(6). With the development of renal biopsy techniques, it has been found that many patients diagnosed clinically with DN may have non-diabetic renal disease (NDRD)(7). This disease may exist alone or coexist with DN. Early detection of non-diabetic renal disease (NDRD) has become critical in our clinical practice. Membranous nephropathy (MN) is a common cause of primary glomerular disease in diabetic patients(8). DN and MN can manifest as proteinuria and impaired renal function, which are difficult to distinguish without renal biopsy(9). Several studies have found that compared with DN patients, NDRD or DN+NDRD patients show significant improvement in proteinuria and renal function after receiving systemic treatment with glucocorticoids, immunosuppressants, antihypertensives, and lipid-lowering drugs(10). However, for DN patients, the use of steroids and other immunosuppressive drugs (used to treat MN) can worsen glycemic control and increase the risk of infection in diabetic patients. Although membranous nephropathy is a commonly occurring primary glomerular disease in non-diabetic individuals, data on its natural progression, treatment, and outcomes in diabetic patients are limited.

Previous magnetic resonance (MR) studies have firmly established a causal link between diabetes and a range of systemic diseases, encompassing gastrointestinal disease, carcinoma psychiatric disorders and so on(11–13). Nevertheless, a comprehensive overview of the association between diabetes and MN remains elusive, hindering our comprehension of severe kidney diseases associated with diabetes. Consequently, we conducted an MR analysis to evaluate the causal relationship between diabetes phenotypes and MN, considering this challenge. MR is an analytical method to infer causal relationships between exposures and outcomes(14). It utilizes instrumental variables, which are single nucleotide polymorphisms (SNPs) identified from genome-wide association studies (GWAS)(15). These SNPs are widely employed as instrumental variables (IVs) because their alleles are randomly assigned and independent of confounding factors such as gender and age and are unaffected by the outcome (e.g., the disease itself). Compared to traditional observational studies, MR methods effectively address issues related to confounding factors and reverse causality. In this study, we conducted MR analysis using phenotype data for diabetes phenotypes and MN obtained from large-scale, openly accessible GWAS data.

## 2 Data source

We extracted the MR analysis data from the IEU Open GWAS database, which predominantly comprises publicly available GWAS summary datasets(Figure 1). The statistical data for membranous nephropathy (MN) was obtained from a GWAS study encompassing 7,979 individuals, of which 2,150 were cases of primary MN and 5,829 served as controls(8). These individuals were drawn from five European cohorts. The diagnosis of idiopathic membranous nephropathy cases was confirmed through the gold standard diagnostic method of renal biopsy. Notably, secondary factors such as patients with hepatitis B virus infection, drug-induced causes, and malignant tumours were excluded from the analysis. Regarding diabetes, fasting glucose, fasting insulin and HbA1C data, we accessed the UK-Biobank, Within family GWAS consortium, MAGIC, Finnish database, MRC-IEU, and Neale Lab, which provided sample sizes ranging from 17,724 to 298,957(16–20). It is essential to mention that all the databases above predominantly comprise individuals of European descent.

**Figure 1:**
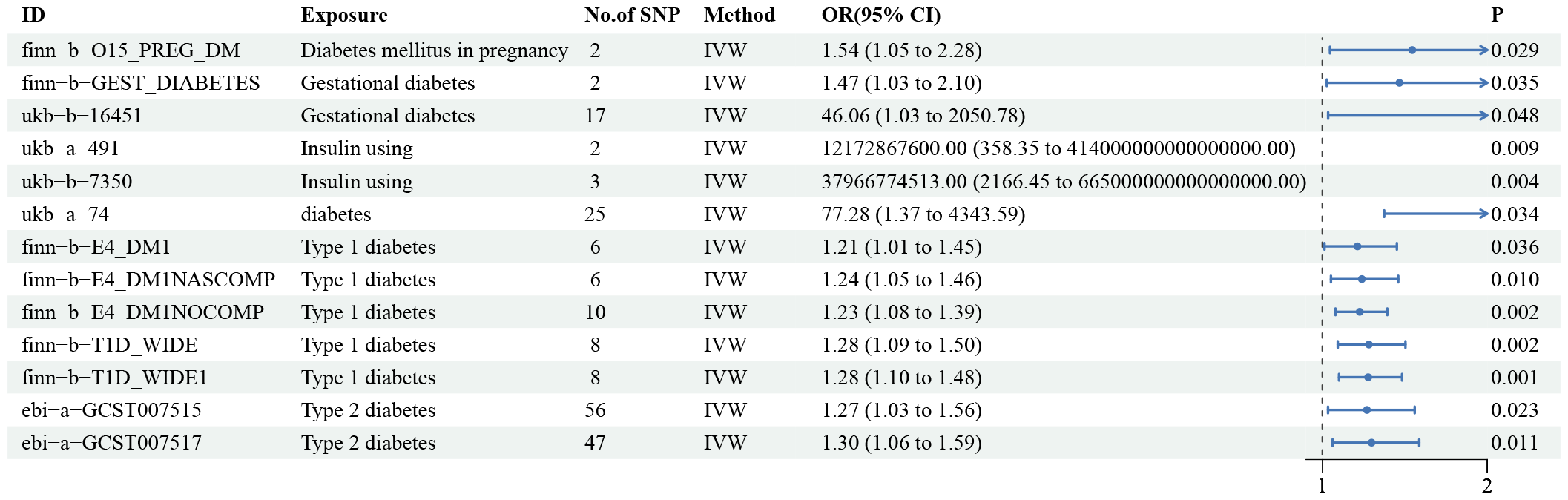
The flowchart demonstrates the experimental design of this study.

## 3 Methods and materials

### 3.1 Selections of Instrument Variants

The instrument variants (IVs) utilized in this MR study must adhere to three fundamental assumptions (Figure 1):

i. IVs should demonstrate strong associations with diabetes phenotypes. IVs with an F statistic less than 10 [F = (β/ SE) ^2^] are considered weak and therefore excluded.
ii. IVs need to be independent. IVs associated with diabetes phenotypes were identified at a genome-wide significance level (P < 5×10^−8^). IVs in high linkage disequilibrium (r^2^) were identified using a European reference panel (1000 gene project)(21), with exclusion criteria of r^2^ > 0.001 or within 10,000 kb.
iii. IVs must be unrelated to the MN, influencing the outcome solely through indirect diabetes traits. Additionally, potential confounding factors that could impact the association between diabetes traits and MN were accounted for. These confounders include hepatitis B, hepatitis C, malignancies, and medications (such as non-steroidal anti-inflammatory drugs, gold compounds, d-penicillamine, bucillamine, and cyclosporine). Any IVs that could potentially act as confounders were removed.

### 3.2 Mendelian randomization

To ensure the robustness of our findings, we employed a range of methods in the MR analysis, including Inverse variance weighted (IVW), MR Egger, Weighted median, Weighted mode, Maximum likelihood, Constrained maximum likelihood (CML), and Penalized weighted median. Among these approaches, IVW analysis emerges as the most effective MR analysis when genetic instrument pleiotropy is absent, and the sample size is sufficiently large(22). IVW estimation consistently provides reliable and close-to-true effect estimates. Consequently, we designated IVW as the primary MR method. In instances where heterogeneity analysis yielded significant results (P < 0.05), we employed the IVW random effects model. The MR Egger method was utilized to examine directional pleiotropy and causal associations, assuming the invalidity of all SNPs as instruments(23). Weighted median analysis was employed when at least half of the SNPs had valid instruments(24). CML evaluated population-wide overlap by maximizing the likelihood function, resulting in lower standard errors. The Penalized weighted median estimator was also employed as a supplementary method, modifying the standard weighted median MR. Causal estimate results from both primary and supplementary MR analyses were visualized using scatter plots, while funnel plots were used to depict the distribution of individual SNP effects. The MR analysis results are presented in the accompanying table.

### 3.3 Sensitivity analysis

The sensitivity analysis comprises heterogeneity tests, pleiotropy tests, and leave-one-out analysis. Our objective is to ensure that the significant results of the MR do not demonstrate heterogeneity or horizontal pleiotropy in the sensitivity analysis. The heterogeneity of SNPs in each analysis is assessed using Cochran’s Q statistic, with P > 0.05 indicating the absence of heterogeneity in the MR results(25). Horizontal pleiotropy may result in false positive findings when instrumental variables are linked to multiple independent phenotypic effects. The MR Egger intercept test assesses horizontal pleiotropy, with P > 0.05 indicating the absence of evidence for horizontal pleiotropy(23). Leave-one-out analysis entails the sequential removal of instrumental variables for reanalysis to evaluate the potential bias of individual SNPs on causal estimates. Since multiple databases are utilized for specific subtypes of diabetes, we will acquire multiple MR results. Since multiple databases are used for certain subtypes of diabetes, we will acquire multiple MR results. In cases where two or more MR estimates for the same outcome derived from non-overlapping samples exist, meta-analysis will be employed to derive a pooled estimate.

### 3.4 Reverse Mendelian randomization analysis

To avoid directional confounders, we will consider MN as the exposure and various subtypes of diabetes as the outcomes for a subsequent round of MR analysis.

### 3.5 Meta-analysis

Due to the duplication of diabetes-related phenotype databases, we conducted MR analysis on phenotypes with multiple databases and obtained various results. We then performed a meta-analysis of the above results using a random-effects model(26).

## 4 Results

The MR analysis revealed significant associations between type 1, type 2, maternal, and gestational diabetes and insulin use in diabetic patients with an elevated risk of MN(Figure 2 and 3, Supplement Tables 1-4). We also found that the genetic prediction of fasting insulin, fasting blood glucose, and HbA1c levels were not associated with the risk of MN(Supplement Tables 5-7). An increment of one standard deviation (SD) in the genetic prediction of diverse diabetes-related phenotypes is associated with a higher odds ratio (OR) for MN. The scatter plots depict the causal relationships and effect sizes for each MR method. The funnel plots exhibit a well-balanced distribution of SNP effects. Leave-one-out analysis identified no influential outliers impacting the final estimates, and the genetic prediction-based causal effects of distinct diabetes subtypes on MN remained consistent with the initial MR analysis even after sequentially removing individual SNPs. This indicates the stability and robustness of our MR study. No evidence of horizontal pleiotropy or heterogeneity was observed (P > 0.05), suggesting that other potential confounding factors are unlikely to introduce bias in our MR study. In the reverse MR analysis, we did not find any evidence of reverse causality(Supplement Table 8). The meta-analysis results of diverse phenotypes across multiple databases, employing a random-effects model, exhibited statistical significance across the board, corroborating our MR analysis findings. This provides further confirmation of the potential heightened risk of MN associated with various diabetes-related phenotypes.

**Figure 2:**
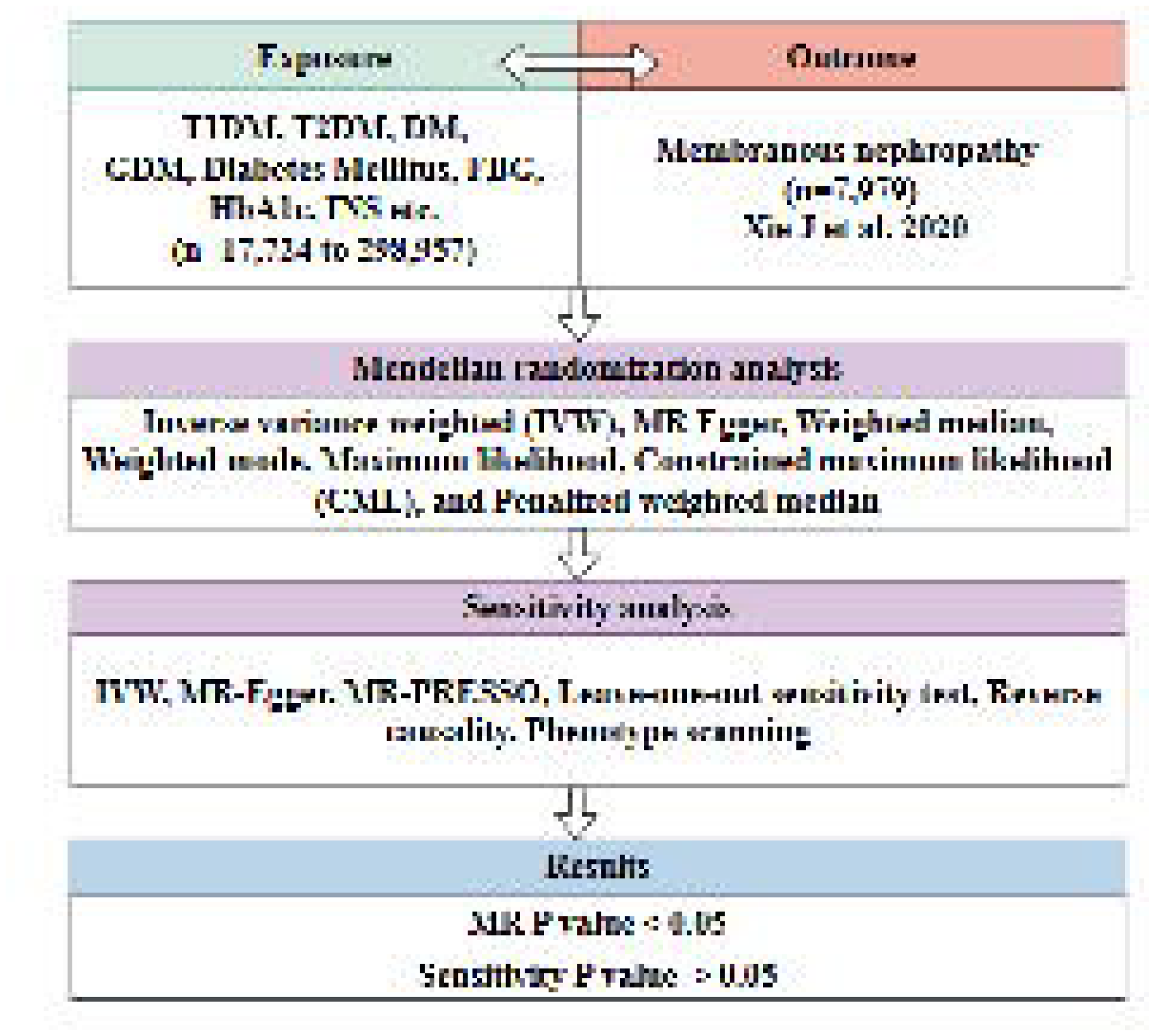
The forest plot shows the results of the MR analysis. OR: odds ratio, IVW: Inverse variance weighted, CI: confidence interval

**Figure 3:**
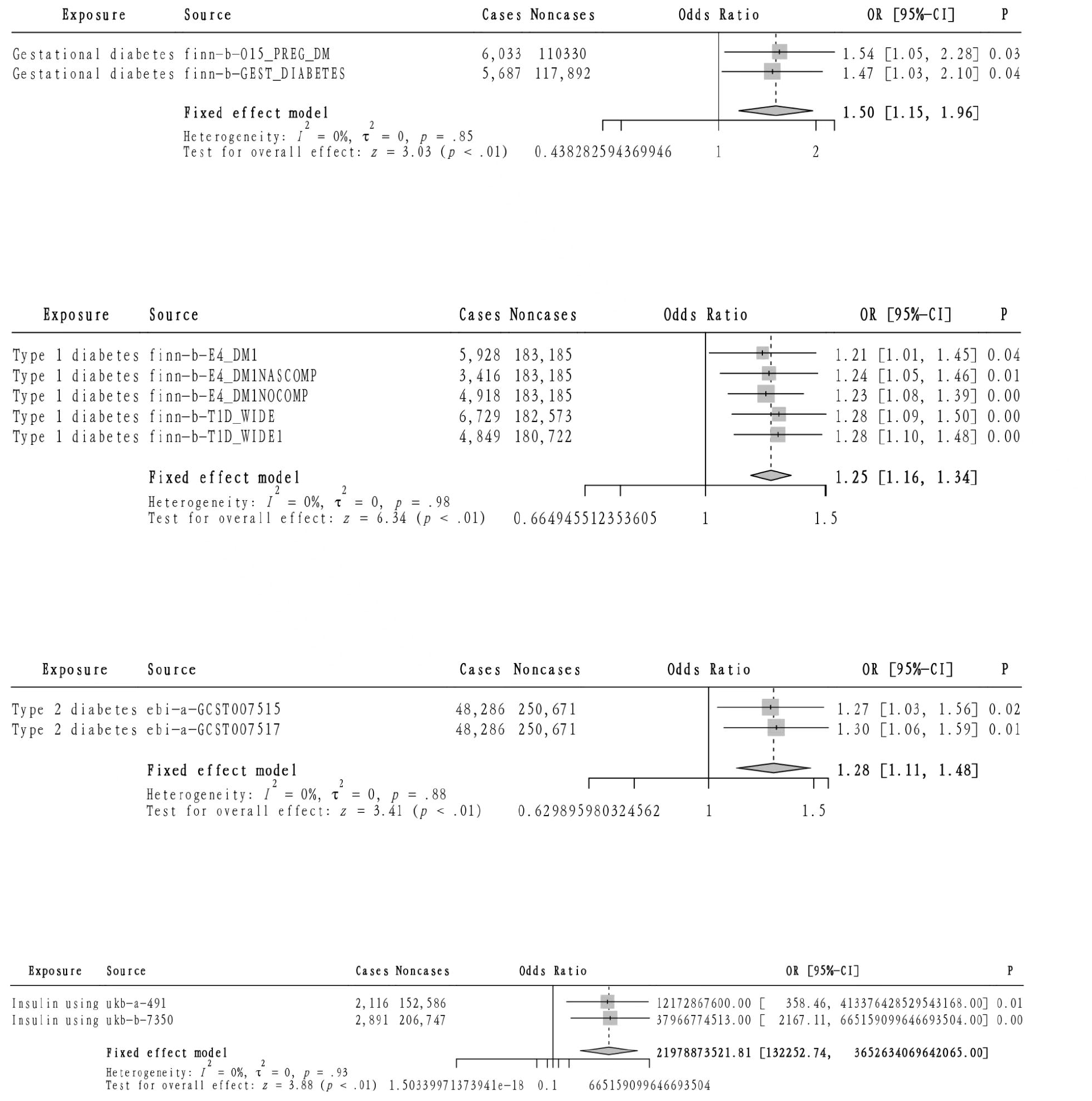
meta-analysis results

## 5 Discussion

The causal relationship between diabetes and MN remains a complex and multifaceted subject. In recent years, the advancement of medical research has led to a gradual recognition of the intimate association between diabetes and kidney diseases, particularly MN. This article aims to delve deeply into the causal link between diabetes and MN, aiming to enhance public awareness and comprehension of these diseases. This study marks the first attempt to integrate GWAS data from extensive cohorts, utilizing multi-sample MR analysis and meta-analysis to assess the causal relationship between various diabetes-related phenotypes and MN. Based on gene prediction, our findings ultimately revealed that type 1 diabetes, type 2 diabetes, gestational diabetes, and insulin use in diabetic patients in diabetic patients were significantly associated with an elevated risk of MN. Sensitivity analysis further corroborated the positive causal relationship between these phenotypes and MN.

Diabetes is a metabolic disease marked by persistently high blood glucose levels, potentially linked to genetic predisposition, environmental factors, autoimmune responses, and other contributing elements. MN, on the other hand, is a glomerular disorder associated with immune complexes. Its primary manifestation is the deposition of these immune complexes on the epithelial side of the glomerular capillary wall, often accompanied by widespread thickening of the basement membrane.

Our study revealed a positive correlation between MN and various diabetes types, including type 1, type 2, gestational diabetes, and insulin usage among diabetes patients. Furthermore, Mendelian randomization studies provide evidence for a causal linkage between genetic predisposition to diabetes and an elevated risk of MN. However, our analysis indicated that genetically predicted levels of fasting insulin, fasting glucose, and glycosylated hemoglobin did not correlate with the risk of MN, suggesting that the association between diabetes subtypes and MN is not primarily mediated by glucose dysregulation. Discovering a link between impaired glucose homeostasis and gastrointestinal diseases holds potential clinical significance. This suggests that apart from managing type 2 diabetes, pharmacological or lifestyle interventions aimed at decreasing circulating glucose levels and fasting insulin levels could potentially contribute to preventing gastrointestinal diseases.

Numerous studies have consistently demonstrated that diabetes mellitus serves as a primary etiology of kidney disease. Among patients diagnosed with diabetes, approximately 30% to 40% develop kidney disease, with MN being a notable manifestation. However, our analysis indicated that the association between diabetes and MN might not be mediated by glycemic dysregulation but rather by other pathophysiological mechanisms. For instance, the immune system function of diabetic patients may be compromised, leading to the deposition of immune complexes on the epithelial side of the glomerular capillary wall, thereby precipitating MN(27). In addition, the incidence of insulin resistance (IR) is continuously increasing in T1DM, T2DM, and gestational diabetes, especially in overweight individuals and patients treated with insulin(28). IR can activate the complement system to induce membranous nephropathy (29). On the other hand, diabetes can cause metabolic disorders such as glucose, protein, fat, water, and electrolytes, promoting inflammation. Lipid accumulation can damage podocytes (a structure and function crucial for maintaining the glomerular filtration barrier of the glomerulus) and induce podocyte degeneration and foot process effacement(30). Podocyte apoptosis is a critical process in the development of MN (30). Moreover, chronic inflammation, including C-reactive protein, mononuclear chemotactic protein-1, interleukin 8, etc, caused by diabetes can stimulate the body to secrete various inflammatory factors, exacerbating kidney damage and glomerular sclerosis(31).

The causal link between diabetes and MN is well-established, yet the mechanism underlying this relationship remains elusive. Current research has yet to fully elucidate the intricate pathways through which diabetes triggers the entire process of MN, leaving this area ripe for further exploration.

It is crucial to acknowledge that, according to genetic predictions, MN does not elevate the risk of various diabetes types. Although MN may result in impaired renal function, it is not directly implicated in the onset of diabetes. Instead, the occurrence of diabetes is influenced by a multitude of factors, including heredity, environmental conditions, and autoimmunity. In contrast, MN primarily represents a kidney disease that may be induced by diabetes. Consequently, for diabetic patients, the prevention and management of MN are paramount. The risk of MN can be significantly mitigated by regulating key parameters such as blood sugar, blood pressure, and blood lipids. Furthermore, for patients who have already developed MN, it is imperative to adopt proactive treatment strategies to decelerate the progression of the disease and safeguard renal function.

This study exhibits numerous strengths. Chief among them is the utilization of the MR methodology, which significantly mitigates the potential for confounding and reverse causality biases. By leveraging summary-level data from extensive genetic studies conducted in European populations, our findings are unlikely to be skewed by population structure biases. Furthermore, estimating these associations across independent data sources and their subsequent combination through meta-analysis enhances the statistical power and the robustness of our research outcomes.

We must also acknowledge several limitations. Firstly, given that this is a non-linear MR analysis, we cannot assess the non-linear association between diabetes and MN using aggregate-level data. Moreover, we cannot discount the possibility that diabetes-related SNPs may affect MN through alternative causal pathways. Notably, while additional MR analysis results, such as Weighted median, Weighted mode, and MR-Egger, no longer exhibit statistically significant associations observed in the primary analysis, this could be attributed to a reduction in method efficacy.

Nevertheless, we rely on IVW, the primary MR method, to observe the direction of these associations, thus supporting the consistency of our findings. Additionally, sensitivity analysis was conducted to validate the consistency of our results further, revealing that our findings are mainly unaffected by heterogeneity and horizontal pleiotropy. Secondly, if the dichotomy of continuous risk factors primarily defines diabetes, the MR analysis of diabetes as an exposed binary phenotype may introduce biases due to the exclusion of restrictive assumptions. However, it is essential to note that the threshold of continuous biomarkers does not solely define diabetes; it can also be determined by ICD codes, drug usage, self-reports, or the threshold of HbA1c and blood glucose levels, as employed in this study. Thirdly, our findings reveal no significant impact of fasting blood glucose, fasting insulin, and HbA1c levels on MN. This observation suggests that the association between diabetes mellitus of various types and MN is not primarily mediated by glucose dysregulation or insulin resistance. Notably, most patients in our study had a long-standing diagnosis of diabetes symptoms. These markers alone are clinically inadequate to assess pancreatic function, necessitating further evaluation. Consequently, our data imply that diabetes-induced MN may have alternative underlying mechanisms. Fourth, despite these findings, the odds ratio (OR) obtained from our MR analysis of diabetes remains unexplained by the direct exposure unit, as previously reported in studies. This limitation may hinder directly comparing our research outcomes with the magnitude reported in observational studies. Fifth, SNPs related to diabetes were sourced from the general population, whereas SNPs about blood glucose traits were derived from a population free of diabetes diagnosis. Consequently, even if diabetic patients are included in the outcome data, the extrapolation of the identified blood glucose traits to the entire population or specifically to diabetic patients remains to be validated in future investigations. Sixth, the exclusive inclusion of European populations in this study restricts the generality of the findings to other ethnic groups, such as those in Asia and Africa. Seventh, discrepancies between genetic scoring and clinical diagnosis of diabetes may give rise to potential misclassification of cases.

In our study, we observed a correlation between the genetic predisposition to diverse forms of diabetes and an elevated risk of MN. Notably, fasting blood glucose levels, fasting insulin concentrations, and HbA1c markers did not significantly influence the risk of MN. Furthermore, diabetes appears to be a contributory factor in the emergence and progression of MN, which serves as a manifestation of diabetic kidney disease. The profound comprehension and exploration of this intricate relationship hold immense importance in preventing and managing diabetes and its associated complications. Additionally, enhancing public knowledge and understanding regarding these diseases is paramount to improving the overall health of individuals. It underscores the vital role of early screening and renal disease prevention among diabetic patients.

## Supporting information

details document

## Data Availability

All data produced in the present study are available upon reasonable request to the authors

## 8 Acknowledgments

Thank you to all the contributors of the publicly available databases used in this study.

## 9 Data Availability Statement

All data are uploaded as supplementary materials.

