## Supplementary material for "Causality between Diabetes and MN: Mendelian randomization and meta-analysis": details document

Supplementary Table 1: MR results

Supplementary Table 2: Pleiotropy analysis results

Supplementary Table 3: Heterogeneity analysis results

Supplementary Table 4: MR-PRESSO analysis results

Supplementary Table 5: MR analysis results of fasting blood glucose and glycosylated hemoglobin

Supplementary Table 6: Heterogeneity analysis results of fasting blood glucose and glycosylated hemoglobin

Supplementary Table 7: Pleiotropy analysis results of fasting blood glucose and glycosylated hemoglobin

Supplementary Table 8: Reverse MR results
